# Variation in Covid-19 Cases Across New York City

**DOI:** 10.1101/2020.05.25.20112797

**Authors:** Awi Federgruen, Sherin Naha

## Abstract

The number of confirmed COVID-19 cases, relative to population size, has varied greatly throughout the United States and even within the same city. In different zip codes in New York City, the epicentre of the epidemic, the number of cases per 100,000 residents has ranged from 437 to 4227, a 1:10 ratio. To guide policy decisions regarding containment and reopening of the economy, schools and other institutions, it is vital to identify the factors that drive this large variation.

This paper reports on a statistical study of incidence variation by zip code across New York City. Among many socio-economic and demographic measures considered, the average household size emerges as the single most important explanatory variable: an increase in *average* household size by one member increases the zip code incidence rate, in our final model specification, by at least 876 cases, 23% of the range of incidence rates, at a 95% confidence level.

The percentage of the population above the age of 65, the percentage below the poverty line, and their interaction term are also strongly positively associated with zip code incidence rates, In terms of ethnic/racial characteristics, the percentages of African Americans, Hispanics and Asians within the population, are significantly associated, but the magnitude of the impact is considerably smaller. (The proportion of Asians within a zip code has a *negative* association.)

These significant associations may be explained by comorbidities, known to be more (less) prevalent among the black and Hispanic (Asian) population segments. In turn, the increased prevalence of these comorbidities among the black and Hispanic population, is, in large part, the result of poorer dietary habits and more limited access to healthcare, themselves driven by lower incomes

Contrary to popular belief, population density, per se, does not have a significantly positive impact. Indeed, population density and zip code incidence rate are *negatively* correlated, with a -33% correlation coefficient.

Our model specification is based on a well-established epidemiologic model that explains the effects of household sizes on R0, the basic reproductive number of an epidemic.

Our findings support implemented and proposed policies to quarantine pre-acute and post-acute patients, as well as nursing home admission policies

## I. Introduction

The world continues to search for a fundamental understanding of the dynamics of the current pandemic. For example, we try to understand why, in the United States (US), as of May 17, 2020, 192,000 COVID-19 cases have been reported in New York City alone, while the country-wide total has been mercifully restricted to 1.5 million, a staggering number, nevertheless. In other words, to date, a city representing 2.5% of the US population accounted for 12.8% of the reported cases. Identifying the main drivers of the disease spread has important implications for the public policies we should implement to contain the current epidemic and mitigate the widely expected “second wave”.

*Population density* is widely believed to be such a main driving force. This theory has some, a priori, intuitive appeal. After all, the number of infections in a given region depends on the basic reproductive number R0, defined as the average number of cases directly generated by a single case, in a population in which all individuals are susceptible. This reproductive number, in turn, depends, *in part*, on the number of individuals with whom a single case has physical contact during the time interval in which he or she is contagious. *Ceteris paribus*, the latter can be expected to be positively correlated with the population density.

However, the theory is challenged, first of all, on an international basis. Many cities with population density as great as or greater than New York City’s 10,198 residents per square kilometre (sq km) have reported much lower absolute and relative case rates. These cities include, for example, Manila (46.128 residents/sq. km), Baghdad (32,874 residents/sq. km), Mumbai (32,303 residents/sq. km), Seoul (16,000 residents/sq. km), Mexico City (9,800 residents/sq. km), and Singapore (8,358 residents/sq. km). Their case incidence rates per 100,000 residents vary between 9.4 (Seoul) and 635.4 (Singapore), as compared to New York City’s rate of 2286 cases per 100,000 residents.

The lower rates in those cities may, possibly, be explained by ex-ante differences in international traffic patterns in and out of the country, affecting the cluster of “imported” cases, or the specific containment and testing policies adopted by the respective governments. However, among states within the US, California has one of the highest population densities (251.3 residents per square mile but one of the lowest COVID mortality rates (8 per 100,000 residents), while Louisiana, with a population density 2.5 times lower than that of California, has reported 49 COVID deaths per 100,000 residents.

And stark differences are apparent within New York City, itself. Wadhera et al. (JAMA, April 29, 2020) reported recently that among the city’s five boroughs, Manhattan had by far the *fewest* hospitalizations per 100,000 residents (Figure 1), but the greatest population density, 2.5 times the citywide average (25,106 vs 10,198 residents/sq. km). (Because the percentage of confirmed cases that require hospitalization is remarkably robust throughout the country, hospitalization rates can be viewed as proxies for incidence rates) In fact, at zip code granularity, the rates of reported cases and the population densities are *negatively* correlated, with a correlation coefficient of – 33% (Table 2).

**Fig1.**
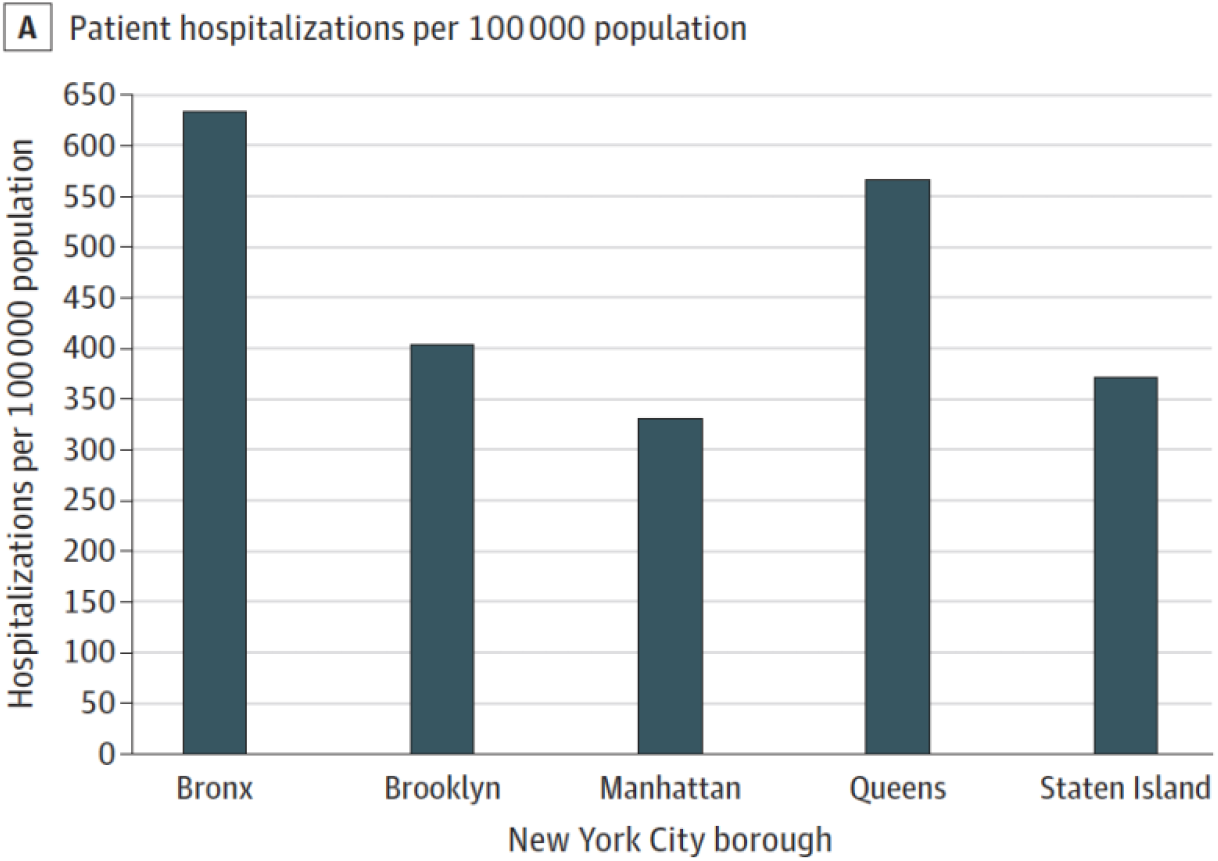
Hospitalization rates due to Coronavirus Disease 2019 (COVID 19), from Wadhera et al. (JAMA, April 29, 2020)

Several authors, including Barr and Tassier (Scientific American, April 17, 2020) and Bassett (New York Times, May 15 2020), have cautioned against relying on *population density* as a prime explanatory variable..

First and foremost, populations consist of “households” and the inter-household and intra-household dynamics of an epidemic differ fundamentally. In particular, the contact rate between a pair of individuals living in the same household tends to be much greater than that between two individuals from different households. Starting with the seminal papers by Bartoszynski (1972) and Becker (1977), this observation has been at the core of many epidemiological models. See Section 2 for a brief description of a seminal model by Becker and Dietz (1995).

The Novel Coronavirus Pneumonia Emergency Response Epidemiology Team in China (China CDC Weekly, 2020) reported that 80% of transmission clusters of the coronavirus occurred *within households*. Nearly three weeks into the initial coronavirus outbreak in Wuhan, China, the government instituted social distancing and travel bans. This had the substantial impact of reducing the reproductive number R0 from 3.88 to 1.26. However, it wasn’t until Wuhan instituted a centralized quarantine where anyone with COVID-like symptoms (e.g. cough, fever, etc.) would be centrally quarantined at hotels or dormitories, that the estimate for R0 was reduced to 0.32, a 75% improvement over social distancing alone! Indeed, this intervention was the final step in achieving full containment of the virus in China.

In New York City, household size varies by zip code, from a minimum of 1.57 to a maximum of 3.97 (Table 1). It is also highly *positively* correlated with the number of confirmed cases per 100,000 residents (Table 2). And although Manhattan is the most densely populated of all New York City boroughs, it is also the one with the *smallest average household size*. Thus, any statistical model intended to explain the variation in case intensities across the City should include the average household size as an explanatory variable.

More broadly, we need to identify demographic and socio-economic indicators that have significant predictive power. New York City is an ideal arena in which to pursue this investigation; as mentioned, the city has, by far, the highest confirmed incidence rate in the country, but it also exhibits remarkable diversity with respect to many potentially relevant demographic and socioeconomic factors. Combining various data sources, described below, we have been able to assess these factors, on a zip-code by zip-code basis. (Complete data were available for 174 of the 177 zip codes in the City.) Table 1 provides a few examples.

**Table 1:** Descriptive statistics by NYC zip code.

|  | **Minimum** | **Maximum** | **Average** |
| --- | --- | --- | --- |
| **# confirmed cases per 100,000 residents** | 436.91 | 4226.51 | 2077.79 |
| **Population density= # residents per sq. mile** | 1389.08 | 126,067.69 | 38 480.75 |
| **Average household size** | 1.57 | 3.97 | 2.65 |
| **Percentage below poverty line** | 2.20 | 45.90 | 16.83 |
| **Percentage above the age of 65** | 0.46 | 28.98 | 14.26 |
| **Percentage of total Hispanic population** | 1.12 | 75.77 | 26.35 |
| **Percentage total Black non-Hispanic population** | 0.37 | 90.51 | 20.06 |
| **Percentage total Black population** | 0.37 | 93.81 | 23.69 |
| **Percentage total Asian population** | 0.07 | 72.62 | 14.78 |

## II. A basic household-based epidemiology model

Becker and Dietz (1995) consider a population with households of varying sizes. Let μ_H_, σ_H_ and *cv_H_ = σ_H_ /μ*_H_ denote the mean, standard deviation and coefficient of variation of the distribution of household sizes, respectively. Let b denote the mean number of infectious contacts that an individual makes with individuals outside her own household during her entire infectious period. An infectious contact is one that is close and intensive enough to result in disease transmission.

Becker and Dietz (1995, p.211) derive the following closed form expression for R0, the basic reproductive number of the epidemic:

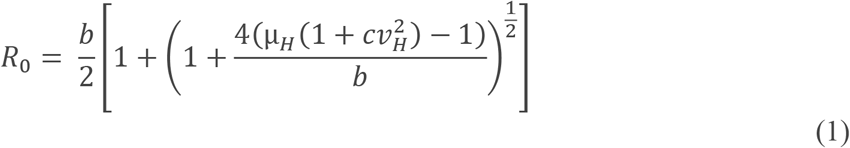

In a population where all individuals live alone, *μH = 1* and *cν_H_ =* 0, so that *R0 = b*, as in elementary models that do not account for household clusters. In contrast, where people live in households of size 4 (e.g., married couples with 2 children), μ*_H_=4* and, if *b >= 1* (i.e., an average infected individual infects at least one individual outside her own household):

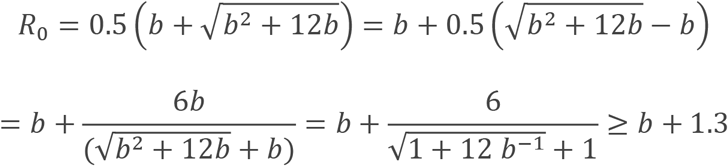

, adding at least a full 1.3 points to the R0 value! And when household sizes vary, as they typically do, the household effect is even greater, see (1). For example, if the household distribution approaches a geometric distribution,

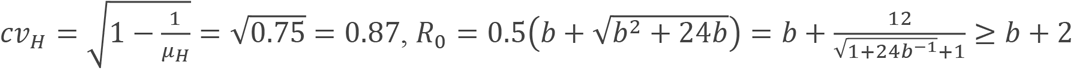

Population density, by itself, may affect R0 via the parameter b, depending on the interactions among individuals in different households. But if it does, (1) shows that it interacts strongly with average household size (and its coefficient of variation) as well.

## III. Methods

The numbers of confirmed COVID19 cases per zip code were obtained from the New York City Department of Health’s GitHub repository. The data are maintained in a CSV (comma separated value) file (see: https://github.com/nychealth/coronavirus-data/blob/master/tests-by-zcta.csv).

Data pertaining to population sizes and population densities per zip code were retrieved from http://zipatlas.com/us/ny/zip-code-comparison/population-density.htm.

All demographic and socio-economic zip code characteristics were extracted from a CSV database created by Buzzfeed, see https://github.com/BuzzFeedNews/2020-05-covid-city-zip-codes/blob/master/data/processed/census/zip_census_data.csv. The database, in turn, was created from 2018 5-year American Community Service (ACS) estimates obtained from the US Census Bureau. The data base contains, in addition to average household size, the following age, economic and ethnic/racial status factors:

Age:

- age_60_and_over
- age_65_and_over
- age_75_and_over
- median_age

Economic:

- pct_more_than_one_occupant_per_room
- pct_below_poverty_level
- household_median_income
- household_mean_income

Racial/Ethnic:

- total_non_hispanic,
- total_white_nonhispanic,
- total_black_nonhispanic,
- total_asian_alone,
- total_black,
- total_hispanic

We applied standard linear regression to estimate best fit regression equations, assuming independent noise terms, all with an identical normal distribution. We have conducted our study in two stages: in the first stage, we focused on crowding, age and income related factors; in the second stage, we investigated to what extent, any of the racial/ethnic characteristics have additional explanatory power.

Based on the empirical and theoretical observations above, we specified a model with (a) *population density* and (b) *average household size* as potential explanatory variables, along with (c) an *interaction term* between them, (d) *the percentage of the population above the age of 65*, and (e) *the percentage below the poverty line*. The relevance of the latter two demographics is both intuitive and explained in our Discussion section. In the remainder of this paper, we employ the following variable names:

- *population_density:* Population density=# residents per sq. mile
- *avg_household_size:* Average household size
- *pct_below_poverty_line:* Percent below poverty line
- *pct_age>65:* Percent above the age of 65
- *cases_per_100k:* # confirmed cases/100,000 residents

As mentioned, the correlation between *population density* and *cases_per_100k* is -33%. The correlation between the standard interaction term *population density*avg_household_size* and *cases_per_100k* is also negative –albeit weaker (−12%). We therefore specified the interaction term as *(population density*avg_household_size)2* which is *positively* correlated with *cases_per_100k*.

**Table 2:** Correlation coefficients among the variables.

|  | Population density= # residents per sq. mile | Average household size | Percent below poverty line | Percent above age 65 | # confirmed cases/100,000 residents |
| --- | --- | --- | --- | --- | --- |
| Population density=# residents per sq mile | 1 | -0.324 | 0.285 | -0.064 | **-0.325** |
| Average household size | -0.324 | 1 | 0.189 | -0.207 | **0.622** |
| Percent below poverty line | 0.285 | 0.189 | 1 | -0.367 | 0.229 |
| Percent above age 65 | -0.064 | -0.207 | -0.367 | 1 | 0.092 |
| # confirmed cases/100,000 residents | **-0.325** | **0.622** | 0.229 | 0.092 | 1 |

## IV. Results

As the correlation figures in Table 2 suggest, average household size is, by far, the single most important explanatory variable in the model. Indeed, it independently explains 62% of the variation in incidence rates. In a model that includes only household size as an explanatory variable, the following equation provides the best fit.

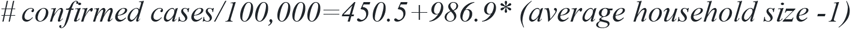

**Table 3:** Regression results with average household size as the single explanatory variable; avg_household size_adj = average household size-1.

|  | Coefficients | Standard Error | t Stat | P-value | Lower 95% | Upper 95% |
| --- | --- | --- | --- | --- | --- | --- |
| **Intercept** | 450.51 | 163.46 | 2.76 | 0.006 | 127.86 | 773.154542 |
| **avg_household_size_adj** | 986.90 | 94.79 | 10.41 | 5.41E-20 | 799.80 | 1174.01 |

The coefficient in front of the average household size variable is highly statistically significant. The equation indicates that in zip code A, where the average household has one more member than that in otherwise comparable zip code B, the case rate would be 986.9 cases/100,000 higher than in zip code B.

However, a significantly superior fit can be obtained by adding the remaining explanatory variables (Table 4). This model accounts for 72% of the variation in case rates (Multiple R=72%), while R2 and the adjusted R2 are 51% and 50%, respectively, resulting in the following regression equation:

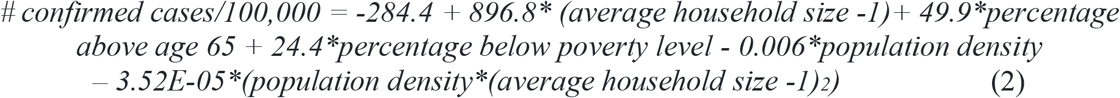

**Table 4:** Full Regression Results without Interaction Effects.

|  | Coefficients | Standard Error | t Stat | P-value | Lower 95% | Upper 95% |
| --- | --- | --- | --- | --- | --- | --- |
| **Intercept** | -284.40 | 353.05 | -0.805 | 0.42 | -981.40 | 412.59 |
| **Population_density** | -0.006 | 0.003 | -1.81 | 0.07 | -0.01 | 0.001 |
| **Average_household_size** | 896.83 | 180.56 | 4.97 | 1.66E-06 | 540.36 | 1253.29 |
| **%_below_poverty_level** | 24.37 | 5.36 | 4.55 | 1.03E-05 | 13.79 | 34.942533 |
| **%_age>65** | 49.94 | 9.80 | 5.10 | 9.29E-07 | 30.59 | 69.2839432 |
| **Population_density\***<br>**(average_household_size)<sup>2</sup>** | -3.52E-05 | 0.001 | -0.03 | 0.98 | -0.002 | 0.00250162 |

The average household size continues to be the main determinant of the number of reported COVID-19 infections. In contrast, neither population density, nor its interaction term with the average household size, has a significant impact on the case intensity. (The intercept value is non-significant as well.)

*Average household size, the percentage of the population above the age of 65 and the percentage below the poverty line*, all have *positive* coefficients that are statistically significant at a very high confidence level. Including the other demographic variables in the model reduces the coefficient of the average household size variable to 896.8 but allows us to conclude at a 95% confidence level, that an increase of but one individual to the average household size in the zip code leads, *ceteris paribus*, to an increase of at least 540.4 cases, per 100,000 residents.

An increase of the percentage of senior residents by one percentage point, augments, *ceteris paribus*, the case rate by approximately 50 cases, more than double the effect of an increase of the poverty rate by a single percentage point. The associations of both demographic factors with zip code incidence rates is plausible;, as well; senior residents may be equally susceptible to getting infected as other age brackets. However, they are at increased risk to develop significant symptoms and complications, thus disproportionally contributing to the counts of *confirmed cases*, the only counts we have, at this point. (A recent study of the population in Santa Clara county by Bendavid et al. (April 30, 2020) estimates that the number of confirmed cases, in that county, may have been as small as 2% of the total number of infected individuals.) Likewise, the *a priori* health status of individuals in the lowest income bracket, is, typically, inferior to that in higher income brackets. Infected individuals with incomes below the poverty line, therefore contribute, disproportionally, to the counts of confirmed cases, as well. Additionally, a much larger percentage of this population segment is employed on location, as opposed to being able to work from home, and is thus more susceptible to infections.

We have checked for the presence of other interaction effects, omitting the population density-related variables that were found to have an insignificant impact. The only significant interaction effect is that between the percentage of the population below the poverty line, and the percentage of senior residents. See Table 5 for the revised regression results. (The addition of the interaction term has a negligible impact on the R2 or adjusted R2 value).

**Table 5.** Full Regression with interaction effect, but population density omitted.

|  | Coefficients | Standard Error | t Stat | P-value | Lower 95% | Upper 95% |
| --- | --- | --- | --- | --- | --- | --- |
| **Intercept** | -549.53 | 246.39 | -2.23 | 0.027 | -1035.93 | -63.14 |
| **avg_household_size_adj** | 1051.69 | 89.00 | 11.82 | 7.17E-24 | 875.99 | 1227.38 |
| **pct_below_poverty_level** | 25.48 | 5.43 | 4.692 | 5.57E-06 | 14.76 | 36.20 |
| **pct age>65** | 48.62 | 9.73 | 4.997 | 1.44E-06 | 29.41 | 67.82 |
| **pct<PL\*pct_age>65** | -0.004 | 0.001 | -3.03 | 0.003 | -0.006 | -0.001 |

Thus, after omitting the population density related variables whose impact was found to be statistically insignificant, our estimate for the coefficient of avg householdsize_adj has a much smaller standard error, resulting in a much narrower 95% confidence interval. The point estimate of this coefficient is now 1051.7, and above 876, at a 95% confidence level. The revised model specification has minimal impact on the estimated coefficients of the renaming explanatory variables.

To visualize the results, we have created an interactive heat map of New York City. The color of each area is determined by the zip code’s *residual* in the above regression equation. This residual denotes the number of cases per 100,000 residents which the zip code experienced in excess of, or below what could be expected based on its demographic characteristics. When clicking on a specific zip code, a table with all of the above measures emerges. See Figure 2 below for a snapshot. The interactive heat map can be accessed from https://colab.research.google.com/drive/1CEY6UadCa3NzwLWoQHPXr_c7dV5llKAU?usp=sharing

**Fig 2.**
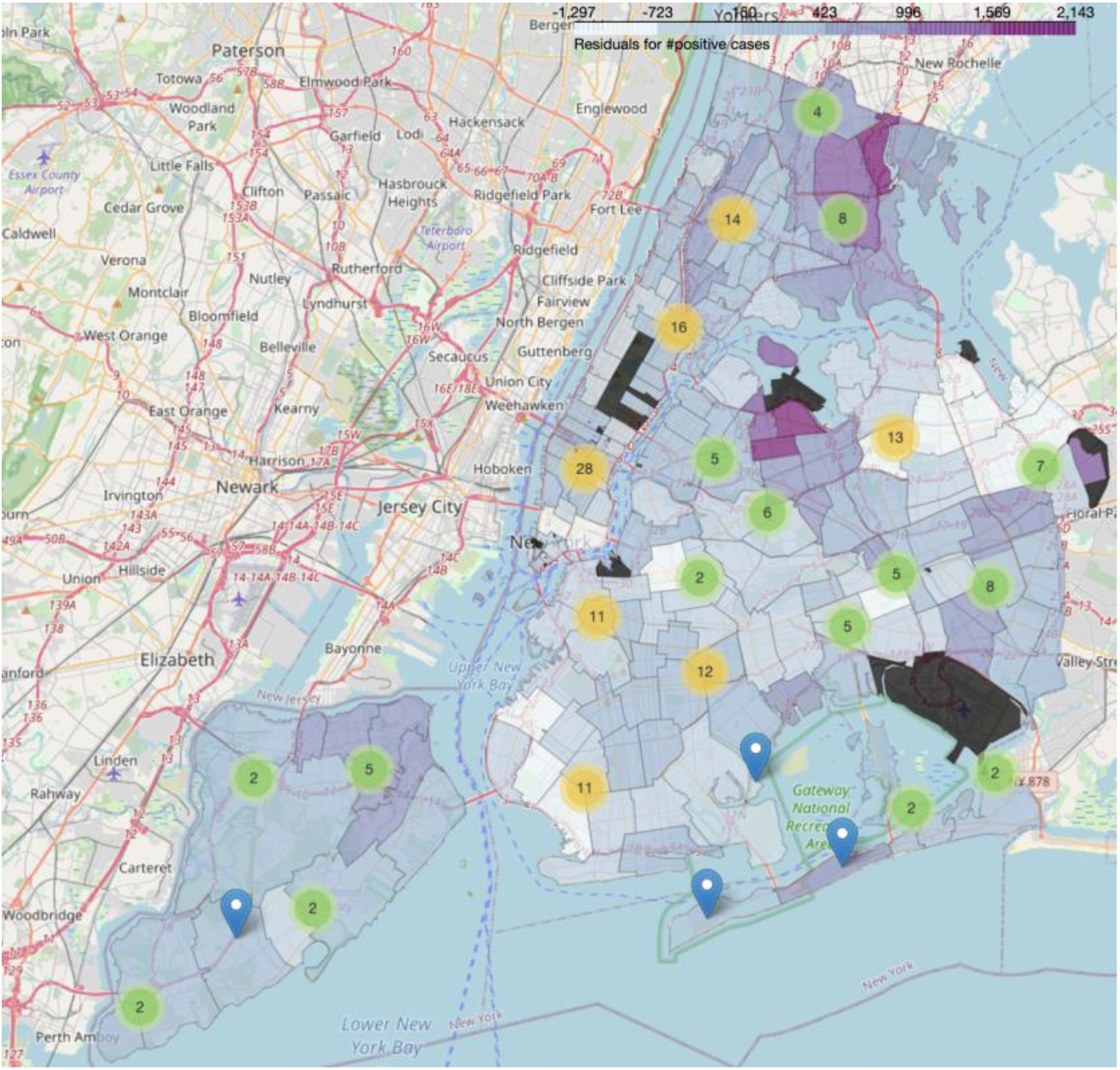
Snapshot of the heat map of New York state showing the zip code’s residuals calculated from regression equation (2).

## IV.1 The impact of racial and ethnic factors

It has been widely conjectured that certain ethnic or racial groups, in particular the black and Hispanic communities, are at increased risk for COVID-19 morbidity. For example, The Illinois Department of Health (April 2020), The Mississippi Department of Health (April 2020) and The Virginia Department of Health (April 2020) have reported that black Americans experienced a disproportionately greater number of reported COVID-19 cases compared to other Americans. In New York City, the black community represents 22% of the population but endured 28% of the confirmed COVID-19 fatalities, see New York State Health Department (April 2020).

However, it remains difficult to assess whether the disproportionately greater incidence rate among the black population, as reported in the above three states, is pervasive throughout the country. The Centers for Disease Control and Prevention (CDC) publishes cumulative COVID-19 data reported by State Health Departments; however, 78% of those data were missing race/ethnicity disaggregation as of April 15, 2020., and 50% as of May 30, see Millett et al. (2020) and Magoy and Wood (2020), the latter reporting on demographic data collected by the COVID Racial Tracker, a joint project of the Antiracist Research & Policy Center and the COVID Tracking Project.

It is easier to relate the case incidence rates in the country’s 3142 counties to the proportions of black residents in these counties, since complete data are available, here. Figure 2 in Millett et al.(2020) shows that the case incidence, mostly, although not universally, increases with the county’s proportion of black residents: on the estimated curve, the case incidence rate doubles when comparing counties with a 1% black population with those where 75% of the population is black

Moreover, even if the higher incidence rate is pervasive, we need to understand whether the difference with the rate among the general population is due to other socio-economic or demographic factors, (such as average household size or the poverty rate), or not.

mitt et al.(2020) provided initial insights. They partitioned the country’s 3142 counties into two sets: the first set of 677 counties with a disproportionately large (> 13%) black population and the second set with the remaining 2465 counties. The estimated case incidence rate in the first set is more than 4 times that in the second, but the associated 95% confidence intervals are overlapping.

Similar concerns apply to the Hispanic/Latino population. With the above proviso that in 48% of confirmed cases the race or ethnicity of the patient is unknown, the COVID Racial Tracker reports that in 42 states plus Washington D.C., Hispanics/Latinos make up a greater share of confirmed cases than their share of the population. In eight states, it’s more than four times greater.

To assess the impact of various ethnicity/racial factors, we have evaluated four additional regression models, each time adding to the model in Eq.(2)-see Table 4_ one of the following four ethnic /racial population percentages as an explanatory variable: (a) total_hispanic, (b)total_black_nonhispanic,, (c) total_black and (d) total_asian_alone,. (We omitted the population density related variables that were shown to have an insignificant impact, see above.).

The Appendix displays the results in Tables 6-9, respectively. The addition of the ethnic/racial descriptors has little impact on the estimates of the coefficients and standard errors of our two main explanatory variables: average household size and the percentage of residents age 65 or older.

The coefficient of the *percentage below the poverty line*, is halved, from approximately 25 to a number in the range [12.27,14.79] in Tables 7-9; In the remaining model (Table 6) where the impact of the proportion of Hispanics is assessed, this coefficient loses its significance. In this case, it is difficult to separate its impacts of the ethnic and the economic variable, as the two are highly positively correlated (65%, see Table 10).

The coefficients of each of the ethic/racial descriptors are significant at a very high confidence level, with p-values below 0.0028, confirming the above described conjectures. However, the magnitude of these coefficients is modest, in particular when compared with *the percentage of seniors* and *the percentage below the poverty line*. An increase of the percentage of Hispanic residents by one percentage point, augments, *ceteris paribus*, the case rate by approximately 12 cases. Thus, if in zip code A, the proportion of Hispanic residents is 20 percentage points higher than that in otherwise comparable zip code B, the case rate would be 247 higher than in zip code B.

The impact of the percentage of *black* residents, and that of *black non-Hispanic* residents, while strongly significant, is even smaller. An increase of this proportion by one percentage point, augments, *ceteris paribus*, the case rate by approximately 6 cases. Thus, if in zip code A, the proportion of black residents is 20 percentage points higher than that in otherwise comparable zip code B, the case rate would be 115 higher than in zip code B.

In contrast, the proportion of Asian residents in a zip code has a (highly significant) *negative* association with the case rate, each additional percentage point *decreasing*, ceteris paribus, the case rate by some 14 cases.^1^

## V. Discussion

Average household size emerges as the single most important demographic variable associated with the large variation in infection rates throughout New York City. Depending on which of the two specifications in Tables 4 and 5 is selected, we estimate that an additional single household member increases the number of cases by 892 or 1051.

This result is quite intuitive. It confirms both the above empirical findings and its theoretical underpinnings, in Section II. Of course, the same contagion potential exists in other settings where a significant number of individuals reside together in the same dwelling, for example dormitories and nursing homes. Girvan and Roy (FREOPP, May 2020) report that, in the USA, 40% of COVID-19 induced deaths occurred in nursing homes and assisted living facilities. Their residents face the simultaneous hazard of living in close proximity to many others, as well as belonging to an age group with significantly increased potential for serious symptoms and hence confirmation of their infections. Similarly, Comas-Herrera et al. (May 3, 2020) document that in Australia, Belgium, Canada, Denmark, France, Germany, Hong Kong, Hungary, Ireland, Israel, Norway, Portugal, Singapore, and Sweden, 49.4 % of reported COVID-19 fatalities took place in nursing homes and related facilities.

The proportion of the population above the age of 65 emerges as a second significant explanatory variable in our study; here we estimate that each additional percentage point contributes 50 cases per 100,000. As explained, the association of the proportion under the poverty line with the outcome, such that each percentage point contributes an estimated 24 cases per 100,000, is also plausible: the health status of this segment of the population is generally poorer, enhancing infections with significant symptoms. Moreover, far more individuals in this income bracket are employed as essential workers with limited opportunities for social distancing; for both reasons, individuals in this segment contribute more than others to the case count. The second factor pertains almost exclusively to individuals below retirement age, a possible explanation for the significantly negative coefficient of the interaction term in the final regression equation (see Table 5).

Our findings about the great importance of household size support such policy initiatives as isolation policies for infected individuals, either immediately upon being identified as infected or post-hospitalization, (see, for example, Chan et al. (Business Insider, April 25 2020). Based on their model in Chan et al.(2020b), the authors show: “that the time to reopening [of the economy] can be shortened by 11%. In addition, assuming symptomatic people are infectious, if 50% of them are quarantined before getting sick enough to go to the hospital, we can reduce the risk of developing severe COVID illness and the time till reopening can be shortened by 86%.” Our findings also support the May 11 decision by New York State to cancel its mandate requiring nursing homes to accept COVID 19 patients.

We have identified that racial/ethnic factors have a significant, albeit more modest, association with case incidence rates above and beyond differences in average household size and the percentage below the poverty line with which they are correlated. Richardson et al.(2020) have identified hypertension, obesity, diabetes and respiratory diseases as the most common comorbidities, in a study involving 5700 COVID patients in the New York City area. These comorbidities are known to be more prevalent among the black and Hispanic population segments, thus providing an explanation for the observed ethnic/racial associations. In turn, increased prevalence of hypertension, diabetes and obesity are, in large part, the result of poor dietary habits and more limited access to healthcare, themselves driven by lower incomes.

Future research should try to identify data on other crowding factors, such as number of individuals residing in nursing homes or dormitories. Although the Center for Disease Control and Prevention (CDC) reports cumulative COVID-19 data as reported by State Health Departments, 78% of those data were missing race/ethnicity dis-aggregations as of April 15, 2020, see

## Data Availability

All data sources have been cited in the document

https://github.com/nychealth/coronavirus-data/blob/master/tests-by-zcta.csv

http://zipatlas.com/us/ny/zip-code-comparison/population-density.htm

https://github.com/BuzzFeedNews/2020-05-covid-city-zip-codes/blob/master/data/processed/census/zip_census_data.csv

## VI. Appendix: The impact of racial/ethnic factors

**Table 6:** Regression results when including %total_hispanic population as an additional independent variable.

|  | Coefficients | Standard Error | t Stat | P-value | Lower 95% | Upper 95% |
| --- | --- | --- | --- | --- | --- | --- |
| Intercept | -607.02 | 239.93 | -2.53 | 0.0123 | -1080.66 | -133.37 |
| avg_household_size_adj | 918.81 | 90.26 | 10.18 | 0.0000 | 740.64 | 1096.99 |
| pct_below_poverty_level | 3.82 | 5.95 | 0.64 | 0.5220 | -7.93 | 15.56 |
| pct age>65 | 54.71 | 9.57 | 5.72 | 0.0000 | 35.83 | 73.60 |
| %total_hispanic | 12.35 | 3.03 | 4.08 | 0.0001 | 6.37 | 18.32 |

**Table 7:** Regression results when including %total_black_nonhispanic population as an additional independent variable.

|  | Coefficients | Standard Error | t Stat | P-value | Lower 95% | Upper 95% |
| --- | --- | --- | --- | --- | --- | --- |
| Intercept | -593.85 | 245.16 | -2.42 | 0.0165 | -1077.82 | -109.87 |
| avg_household_size_adj | 961.76 | 90.63 | 10.61 | 0.0000 | 782.84 | 1140.67 |
| pct_below_poverty_level | 14.79 | 5.01 | 2.95 | 0.0036 | 4.89 | 24.69 |
| pct age>65 | 50.42 | 9.71 | 5.19 | 0.0000 | 31.26 | 69.59 |
| %total_black_nonhispanic | 5.87 | 1.94 | 3.03 | 0.0028 | 2.05 | 9.70 |

**Table 8:** Regression results when including %total_black population as an additional independent variable.

|  | Coefficients | Standard Error | t Stat | P-value | Lower 95% | Upper 95% |
| --- | --- | --- | --- | --- | --- | --- |
| Intercept | -600.22 | 244.74 | -2.45 | 0.0152 | -1083.37 | -117.08 |
| avg_household_size_adj | 961.57 | 90.44 | 10.63 | 0.0000 | 783.04 | 1140.11 |
| pct_below_poverty_level | 13.79 | 5.08 | 2.71 | 0.0073 | 3.76 | 23.83 |
| pct age>65 | 50.75 | 9.70 | 5.23 | 0.0000 | 31.61 | 69.89 |
| %total_black | 5.77 | 1.86 | 3.10 | 0.0022 | 2.10 | 9.43 |

**Table 9:** Regression results when including %total_asian population as an additional independent variable.

|  | Coefficients | Standard Error | t Stat | P-value | Lower 95% | Upper 95% |
| --- | --- | --- | --- | --- | --- | --- |
| Intercept | -484.66 | 238.91 | -2.03 | 0.0441 | -956.29 | -13.02 |
| avg_household_size_adj | 1081.07 | 86.49 | 12.50 | 0.0000 | 910.34 | 1251.80 |
| pct_below_poverty_level | 12.27 | 4.89 | 2.51 | 0.0131 | 2.61 | 21.93 |
| pct age>65 | 55.48 | 9.45 | 5.87 | 0.0000 | 36.82 | 74.13 |
| %total_asian_alone | -14.73 | 3.17 | -4.64 | 0.0000 | -20.99 | -8.47 |

**Table 10:**
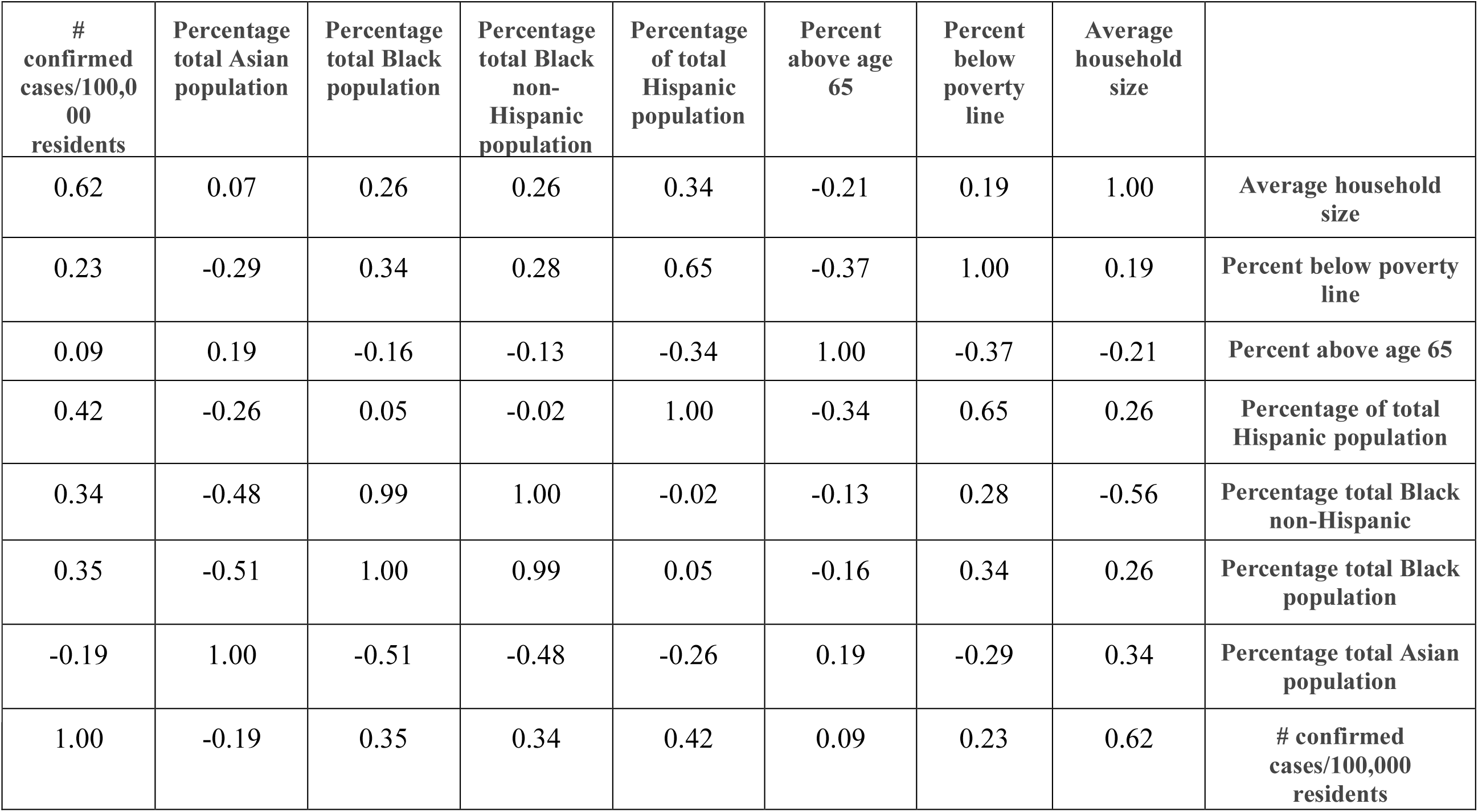
Correlation coefficients among different variables.

## Acknowledgement

We express our gratitude to Dr. Daniel Berman, Infectious Disease Specialist at Montefiore Hospital, Professor Jing Dong of the Columbia Business School, Judith Jacobson, Professor of Clinical Epidemiology at the Columbia School of Public Health, Professor Edward Kaplan of the Yale School of Management and Professor of Public Health at Yale, as well as Professor Assaf Zeevi of the Columbia Business School for their insightful suggestions and comments with respect to earlier drafts of this paper.*Awi Federgruen* is the Charles E. Exley Professor of Management at the Graduate School of Business at Columbia University.

*Sherin Naha* is a Graduate Student in the Management Science & Engineering Masters Program at Columbia University

## Footnotes

1 Ow. The results thus partially confirm and partially challenges conjectures offered by various scholars, e.g., Kendi (2020): “Does this mean Latinos and Asians are being infected with, and dying from, COVID-19 at higher rates than other New Yorkers? We don’t know for certain, but it sure seems that way.”

## Notes

### Competing Interest Statement

The authors have declared no competing interest.

### Funding Statement

No external funding was received

